# Long time frames to detect the impact of changing COVID-19 control measures

**DOI:** 10.1101/2020.06.14.20131177

**Authors:** Jessica E Stockdale, Renny Doig, Joosung Min, Nicola Mulberry, Liangliang Wang, Lloyd T Elliott, Caroline Colijn

## Abstract

**Background:** Many countries have implemented population-wide interventions such as physical distancing measures, in efforts to control COVID-19. The extent and success of such measures has varied. Many jurisdictions with declines in reported COVID-19 cases are moving to relax measures, while others are continuing to intensify efforts to reduce transmission.

**Aim:** We aim to determine the time frame between a change in COVID-19 measures at the population level and the observable impact of such a change on cases.

**Methods:** We examine how long it takes for there to be a substantial difference between the cases that occur following a change in control measures and those that would have occurred at baseline. We then examine how long it takes to detect a difference, given delays and noise in reported cases. We use changes in population-level (*e*.*g*., distancing) control measures informed by data and estimates from British Columbia, Canada.

**Results:** We find that the time frames are long: it takes three weeks or more before we might expect a substantial difference in cases given a change in population-level COVID-19 control, and it takes slightly longer to detect the impacts of the change. The time frames are shorter (11-15 days) for dramatic changes in control, and they are impacted by noise and delays in the testing and reporting process, with delays reaching up to 25-40 days.

**Conclusion:** The time until a change in broad control measures has an observed impact is longer than is typically understood, and is longer than the mean incubation period (time between exposure than onset) and the often used 14 day time period. Policy makers and public health planners should consider this when assessing the impact of policy change, and efforts should be made to develop rapid, consistent real-time COVID-19 surveillance.

## 1 Introduction

In response to the COVID-19 global pandemic, many countries have implemented large-scale physical (“social”) distancing measures. The details of physical distancing vary significantly between and within countries. While government-mandated lockdowns have been enforced in many parts of the world, certain regions have introduced less strict distancing measures. Such differences in the severity and timeliness of the response, as well as variation in patterns of social contact within a community and public compliance, all modulate the effect of distancing measures on a region’s epidemic trajectory. The relaxation of physical distancing must be done gradually and with care. Accordingly, countries experiencing increasing case counts have considered, and will continue to need to consider, what degree of distancing measures to implement in order to reduce disease spread whilst maintaining public compliance and minimising negative effects of distancing (such as adverse health and economic impacts). Understanding the possible trajectories which may arise from changing physical distancing measures is crucial to ensuring that sufficient measures are still taken after relaxation begins. Quantifying the effect and time scale of such relaxations will contribute to health care capacity forecasting, and delivery of timely advice and instructions to communities.

A number of recent studies have examined the effectiveness of government control measures on the spread of COVID-19 [2, 8, 34, 35, 23, 32, 18, 9]. One approach has been to retrospectively compare observed data to baseline model output [34, 8]. Many studies have also focused on estimating changes in the effective reproduction number over time [11, 6, 35]. Similarly, [2] directly estimates the impact of control measures on COVID-19 transmission patterns using a Bayesian model with explicit physical distancing. However, these studies do not focus on reporting the time until a given amount of change is predicted between many simulations of future case counts. Instead, they compare simulations to a constant baseline, or report quantitative measures on only a few simulations.

In this work, we use a likelihood-based approach to determine when we may expect to see the effects of implementing or relaxing physical distancing measures using case count data. This is in contrast to the work described above which focuses on quantifying the effect of such measures. We further use this method to estimate the changes in growth rate that would be expected after beginning to strengthen or relax physical distancing. We use the SEIR-type model developed in [2] to simulate scenarios over a broad range of simulation parameters and strengths of distancing. We consider the impact on the time to detect a change in distancing of data collection delays and inconsistencies, and apply our methods to publicly available case count data from British Columbia, Canada. While we focus here on the “first” implementation and relaxation of physical distancing for COVID-19, these methods can also be used to detect the first time we would expect to see a change in reported case counts in response to modifications of any non-pharmaceutical intervention (NPI). For example, we could use this model to explore the effects of introducing digital contact tracing or improved testing.

## 2 Methods

### 2.1 Transmission model

We use a compartmental model describing susceptible, exposed, infectious and removed individuals (SEIR) [2]. We fit the model to data on daily case counts, and incorporate knowledge of the incubation period and duration of the infectious period in the model. The model includes a fixed proportion of the population who are willing and able to practise physical distancing, although individuals can transition between distancing and non-distancing modes. Upon infectious contact, individuals move from a susceptible state (*S*) to an exposed state (*E*_1_). From here, the disease progresses through the infectious, pre-symptomatic state (*E*_2_), to infectious and symptomatic (*I*), and finally to removed (*R*). Removed individuals are assumed to be not susceptible. The model furthermore incorporates a possible quarantined state (*Q*) following symptom onset, in which individuals are still infected but are unable to infect others. For each state, there is a corresponding “distanced” state. A schematic for this model is shown in Figure 1.

**Figure 1:**
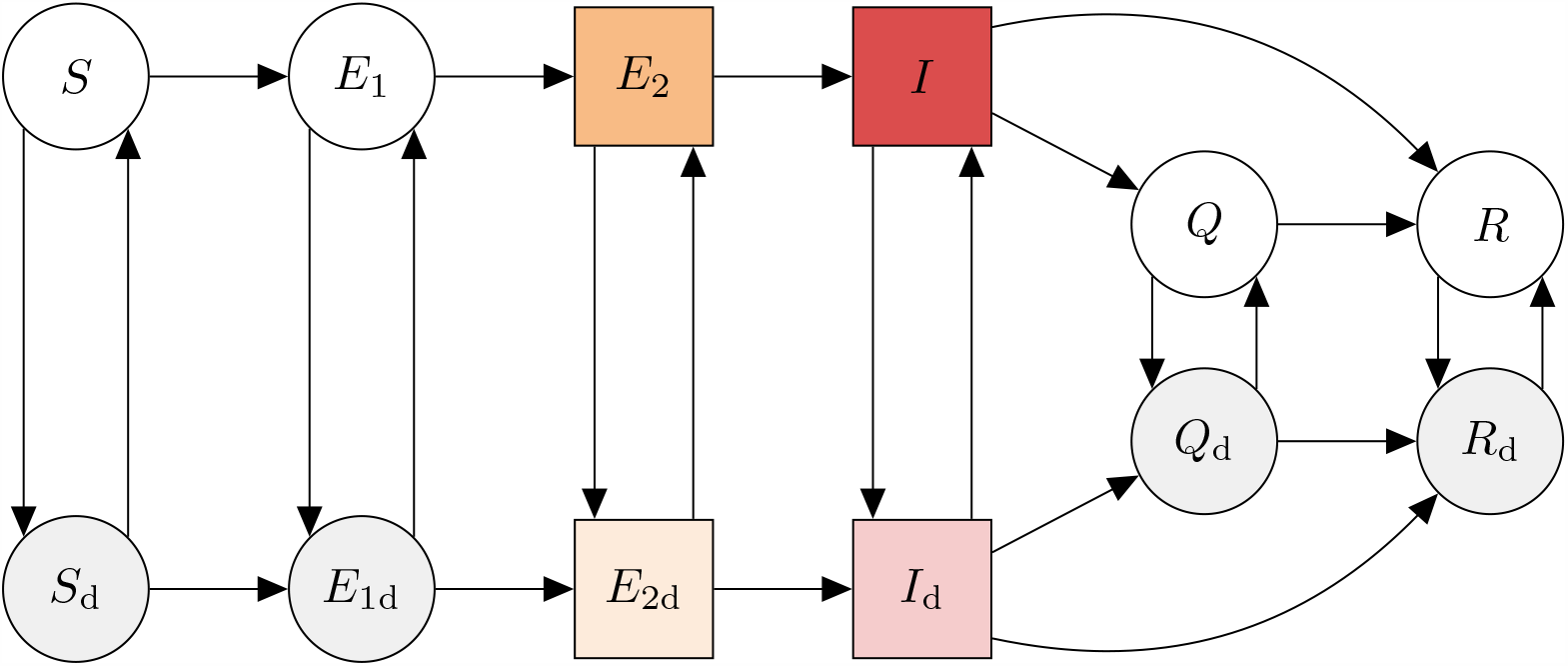
Schematic of the epidemiological model from [2]. Compartments are: susceptible to the virus (*S*); exposed (*E*_1_); exposed, pre-symptomatic, and infectious (*E*_2_); symptomatic and infectious (*I*); quarantined (*Q*); and removed (recovered or deceased; *R*). There are analogous states for individuals practising physical distancing (bottom row). An individual in state *X* can begin distancing and move to the corresponding distanced state *X*_d_ at rate *u*_d_. The reverse transition occurs at rate *u*_*r*_. The model quickly settles on a fraction *e* = *u*_*d*_*/*(*u*_*d*_ + *u*_*r*_) participating in distancing, and dynamics depend on this fraction, rather than on the rates *u*_*d*_ and *u*_*r*_. The red and tinted red squares represent the compartments that we identify as active cases. The orange/orange tinted and red/red tinted compartments indicate infectious cases.

The differential equations for the non-distancing compartments are given as follows:

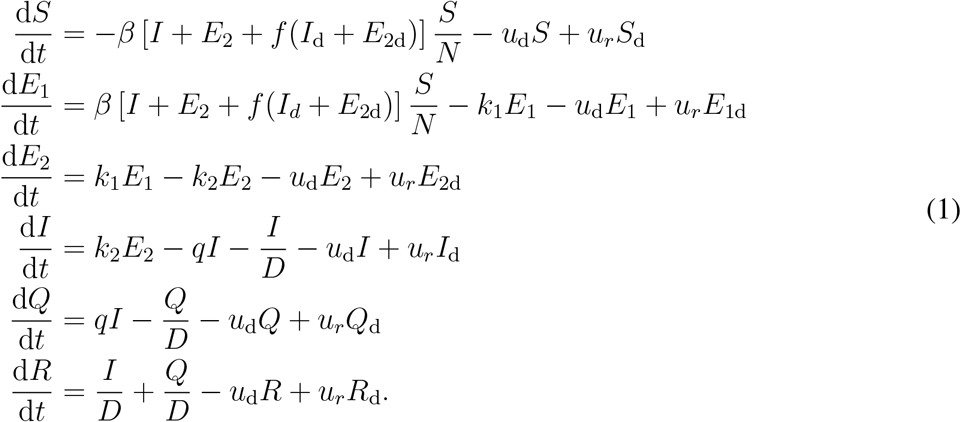

The differential equations for individuals practising physical distancing are analogous:

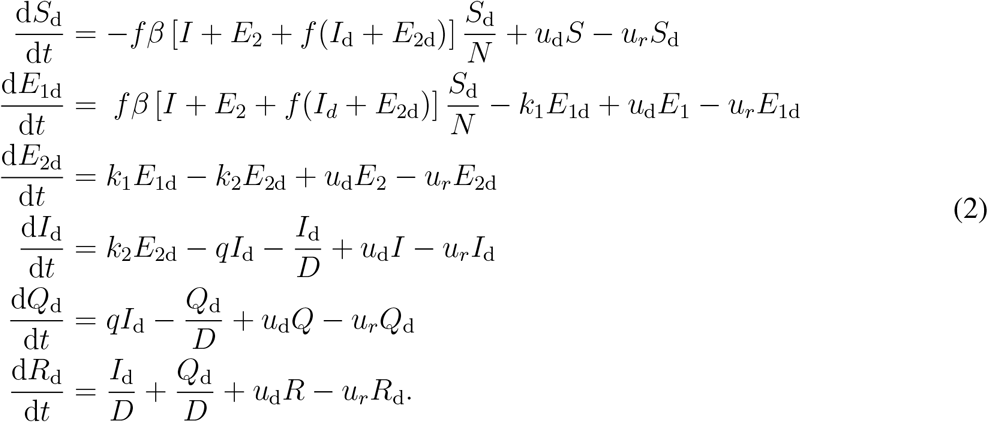

In these equations, the strength of physical distancing is represented by the parameter 0 *≤ f* (*t*) *≤* 1, with *f* (*t*) = 1 indicating no physical distancing. This parameter modulates transmission within the distancing compartments. In addition, individuals in the distancing compartments are less likely to be encountered by others, and so contribute a reduced portion of the force of infection as well. The value of *f* may be allowed to vary in time as physical distancing is strengthened or relaxed in the population. In particular, for the purpose of this work we define:

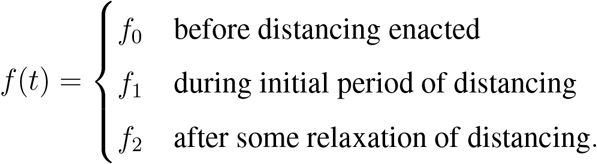

We take the mean number of new cases reported on day *t, µ*_*t*_, to be a weighted sum of those who became symptomatic at some time *s* days ago:

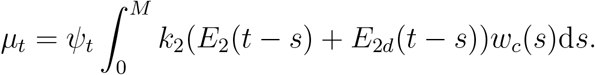

Here *w*_*c*_ represents the distribution of delay between the onset of symptoms and a positive test being reported and *ψ*_*t*_ is the fraction of eligible cases on day *t* that will be tested and reported. The upper limit on the integral, *M*, represents the maximum possible delay in days between onset and reporting, for which we use twice the mean delay. We seek to determine how soon changes in distancing measures will cause substantial changes in the number of prevalent cases. And then, determine how well we may be able to estimate the strength of these measures given the delay and noise in the reported case counts and uncertainty about the model parameters. Given a set of daily case count data {*C*_*t*_}, where *C*_*t*_ is the number of cases identified on day *t*, we use a negative binomial likelihood with mean *µ*_*t*_ and dispersion parameter *ϕ*, NB(*C*_*t*_ | *µ*_*t*_, *ϕ*) as in [2], to write the likelihood of the data given the model parameters accounting for dispersion:

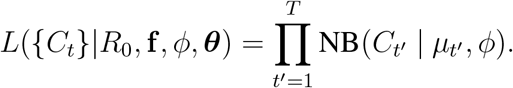

Here *T* is the final day for which case counts are available (or, of concern), f = (*f*_0_, *f*_1_, *f*_2_), and ***θ*** = (*u*_d_, *u*_*r*_, *q, D, k*_1_, *k*_2_, *ψ*_*t*_) are the remaining model parameters, which are assumed to be fixed in all of our experiments except for the simulations in Figure S6. In this parameterization of the negative binomial distribution, the variance scales with the mean according to the dispersion parameter *ϕ*: Var[{*C*_*t*_}] = *µ* + *µ*^2^*/ϕ*. Values of the model parameters for our analysis using British Columbia case count data are provided in Table S1.

### 2.2 Time to pairwise model divergence

The ODEs described above require parameters which can be sampled from prior distributions. In particular, the parameter *R*_0_, the basic reproduction number, is a fundamental determinant of such models’ dynamics. In the model above, 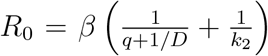in the absence of distancing (*i*.*e*., when *f* = 1). We use uncertainty in *R*_0_ to capture uncertainty in transmission and duration of infection simultaneously: given information about the duration, the transmission parameter *β* can simply be scaled in order to match the exponential growth phase of a dataset. The ODEs can be simulated numerically, drawing *R*_0_ from the prior and keeping other parameters fixed. This procedure induces a distribution over the case counts seen on each day.

We introduce a method for determining the first date at which the case counts of two models begin to diverge. The simulated datasets for a model are treated as random samples from a distribution of prevalence in each component of the ODE. In order to determine when the effect of physical distancing first appears in a model (*i*.*e*., determining when a model diverges from a baseline model), we compute the empirical probability that there is a difference between the two models, according to the distributions of prevalence implied by the two models. In this work, we mainly consider a difference of 10 prevalent and non-quarantined cases, which correspond to *I* +*I*_*d*_ in the model, to be a substantial difference, as this may be a number for which the risk of a return to community spreading is imminent. We refer to this threshold as the number of *active cases* (*i*.*e*., the *I* + *I*_*d*_ component is the number of *active cases*). To compute the empirical probability that a substantial difference occurs on a given day after introducing distancing measures, we consider the proportion of simulated samples for which the model with physical distancing shows fewer infected counts than the non-distancing model by at least 10. (Alternatively, if distancing measures are being relaxed rather than introduced, then prevalent cases would increase and we consider the proportion of simulated samples for which the model *without* physical distancing shows more than 10 active cases.) We form this empirical probability for each day. The first day on which the empirical probability of observing a 10-case difference is at least 0.95 is our estimate for the first day at which the effect of modifying physical distancing is substantial (this is the *days until threshold* in our figures). In most of our experiments involving this threshold, we use a value of 10 for the threshold. In addition, we conduct an experiment in which the threshold is varied in the set {5, 10, 15, 20} for various relaxation parameters.

In predictive investigations of the effects of relaxing physical distancing, we simulate the SEIR-type model displayed in Section 2.1 forwards in time, starting from May 1st and ending on July 1st. We investigate situations in which the amount of physical distancing is modulated on May 17th (the strategy for restarting British Columbia through relaxation of physical distancing recommendations began in the middle of May [28]) through a forcing of the ODE parameter *f*_2_, and in which many of the ODE parameters are modulated through grid-searches. We also consider a retrospective analysis of the introduction of physical distancing in British Columbia, assuming that physical distancing is introduced in mid-March (March 18th) by modulation of parameter *f* from *f*_0_ = 1 to *f*_1_ = 0.4 and *f*_1_ = 0.7, and compare to a baseline model in which *f* remains at 1. We investigate 25% to 50% confidence intervals around the number of active case in early/mid-April, by considering 100 replicates in the ODE simulations, each with a value of *R*_0_ drawn independently from the prior.

In Section 2.3, we provide some evidence that the physical distancing undertaken by people in British Columbia corresponds to a modeled value for *f*_1_ of 0.36. We consider a baseline model in which the value of *f* remains at 0.36 for the duration of the time. We consider alternative relaxation models in which the value of *f* changes from *f*_1_ = 0.36 to either *f*_2_ = 0.5 (weak relaxation), *f*_2_ = 0.65 (medium relaxation) or *f*_2_ = 0.9 (strong relaxation) on May 17th. For each condition, we again consider 100 replicates each with a value of *R*_0_ drawn from the prior.

### 2.3 Time to observe a change in the strength of distancing

In addition to estimating the first time at which we would expect a substantial difference in incidence to occur after a change in physical distancing, we also aim to understand the time it would take to observe such a difference in reported case counts. We use a likelihood-based approach to estimate if a change in distancing measures is yet having an effect on observed case counts, and to estimate the size of this effect.

The likelihood expression, as described in Section 2.1, relates predicted case counts from the SEIR-type model to reported case counts, over time and corrected for the delay between symptom-onset and reporting which is assumed to have a Weibull distribution (*w*_*c*_). This likelihood, written as a function of a given strength of physical distancing *f*_*X*_, (*e*.*g*., *f*_1_, the strength of distancing when it is first implemented or *f*_2_, the strength of distancing after relaxation) may be maximised to find 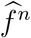, the maximum likelihood estimate (MLE) of *f*_*X*_ on day *n*. Estimation of 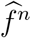 uses data from the start of the outbreak up to and including day *n*. Since case count data can be quite noisy, there can also be considerable variation in the day-by-day MLEs. For example, in the first few days after a change in physical distancing, a single day with a low number of identified cases can have a large impact on the MLE for *f*_*X*_. To mitigate the effect of this noise, we introduce a stopping rule in which an estimate 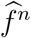 is “accepted” once the MLE changes by less than 5% over a 3-day period. We use this day-by-day MLE approach to estimate the value of *f*_1_ in the period following the introduction of distancing recommendations in British Columbia, using daily case counts reported between March 1st and April 22nd, 2020. We find a credible band by simulating the SEIR model with 50 different values of the baseline *R*_0_ parameter, normally distributed about the mean *R*_0_ = 2.57 estimated by a pre-distancing model fit, and with standard deviation 0.05.

Although, as of the time of writing, we have only started to see the effects of relaxing physical distancing in British Columbia, we simulate the effects of relaxing distancing to 65% of the normal (unrelaxed) level, under the assumption that the noise and delay in observing case counts remains as during March/April in British Columbia (*ϕ* = 5, delay shape parameter 9.85, delay scale parameter 1.73 as estimated in [2]). We also consider other levels of distancing relaxation (50%, 90%) and record the resulting impact on the number of days *n* to accept the MLE 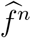. For the credible bands, we increased the standard deviation to 0.15 to obtain a similar level of variation in the model fit as compared to the introduction-of-distancing estimation (the majority of individuals distancing causes the overall reproduction number to be lower).

We also explore the impact of changing the noise/delay of case count reporting on the time to detect a change in the strength of physical distancing. We simulate outbreak trajectories for strengthening distancing under varying levels of the dispersion parameter *ϕ*. In this investigation, we control the variance in the number of daily reported cases: note that this is *not* the same as the dispersion parameter about *R*_0_, commonly referred to as *k*. We also control the shape and scale parameters of the onset-to-reporting Weibull distribution. We then perform the same daily MLE estimation procedure on these simulated time series. Note that this back-estimation requires the assumption that the true noise level and delay are known. If we did not have estimates of these quantities in reality, we would need to co-estimate them along with the strength of distancing, which may introduce identifiability issues or a longer time to acceptance.

All statistical analysis was performed in *R* [24], *and datasets and R* code are available^*^ on *GitHub* under an open source license.

## 3 Results

We find that it takes 21-23 days (March 18th until April 9th or 11th) before there is a substantial difference between the baseline trajectory (the exponential growth phase of the SEIR-type model) and a trajectory with distancing measures of *f*_1_ = 0.4, 0.7. And it takes 26 days to estimate the strength of distancing using reported data. The situation is similar for relaxing measures: it takes approximately 45 days before a substantial difference arises due to relaxing distancing from *f*_1_ = 0.36 to *f*_2_ = 0.65, and 23 days to detect such a difference using observed cases. Figure 2 shows rising case trajectories for which distancing is introduced, and declining ones in which distancing is relaxed, and illustrates the dependence of the timing on the severity of the change. If the change is weak (*e*.*g*., a weak relaxation from *f*_1_ = 0.36 to *f*_2_ = 0.5) there is no discernible difference between the trajectories before July 1 (47 days).

**Figure 2:**
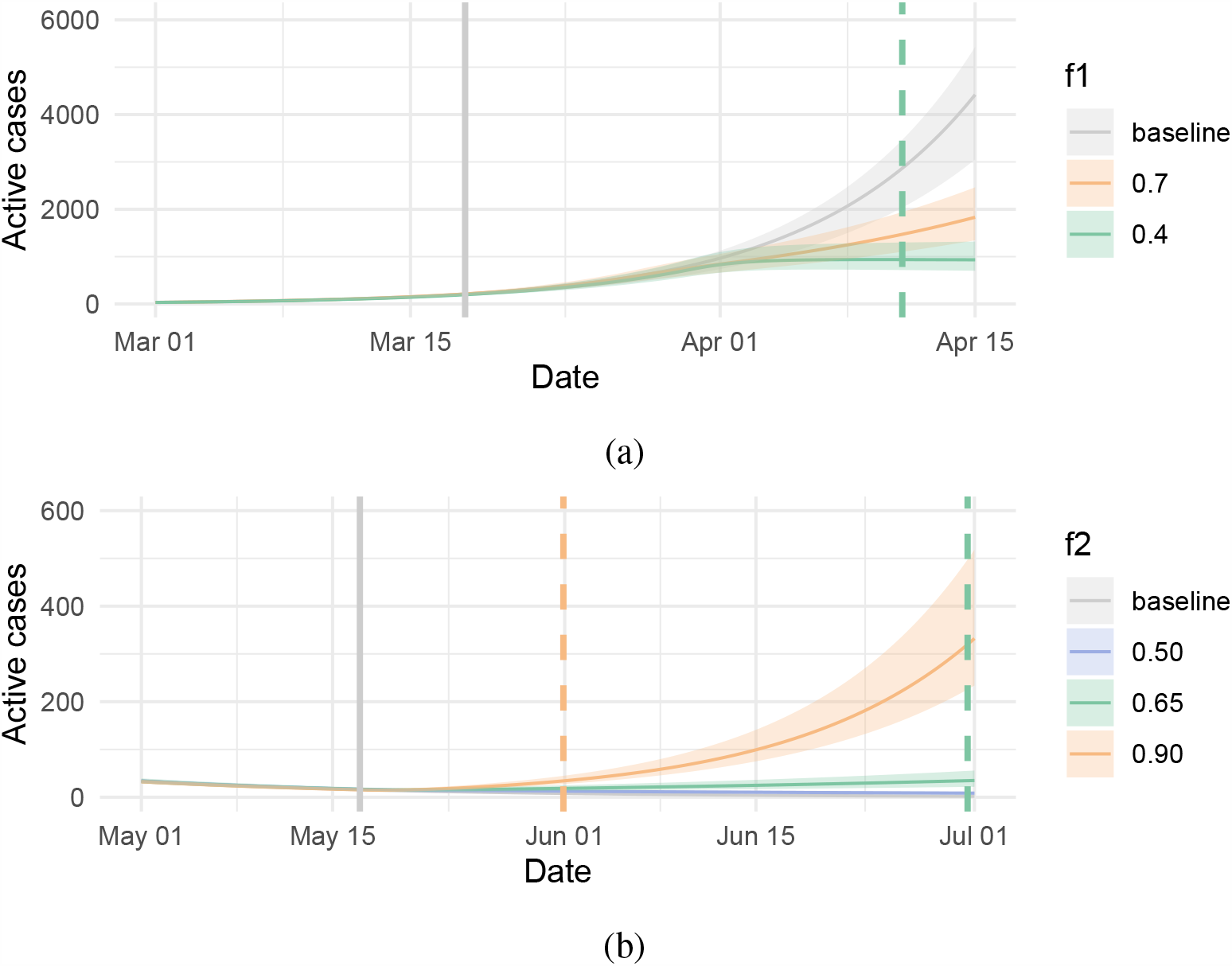
Projected active cases from modulation of physical distancing. (a) An examination of the effects of enacting physical distancing in British Columbia in Spring 2020 (March 18). Three models are considered: A baseline model (grey) in which physical distancing is not introduced, and also two models in which the amount of physical distancing is modified to be *f*_1_ = 0.7 (green line), or *f*_1_ = 0.4 (orange line). Vertical dashed lines indicate when there is a substantial difference between the baseline and the altered trajectory. For *f*_1_ = 0.4 a significant change is identified after 24 days. (b) Projection of the number of active cases (*I* + *I*_*d*_) after relaxation of physical distancing. Three levels of relaxation are considered corresponding to *f*_2_ = 0.5, *f*_2_ = 0.65, or *f*_2_ = 0.9. Dashed vertical lines indicate the day for which *I* + *I*_*d*_ exceeds what is expected under no relaxation by 10 cases. For the relaxation level *f*_2_ = 0.5, no difference is seen for the period under question (the curves for the *f*_2_ = 0.5 and baseline conditions broadly overlap). For *f*_2_ = 0.65 or *f*_2_ = 0.9, significant changes are seen after 44.5 and 14.9 days, respectively.

The time until a substantial difference arises naturally depends on what is considered a substantial difference, and on the underlying uncertainty. We explore the impact of the threshold choice and strength of relaxation (Figure 3); naturally, stronger relaxation (all the way to baseline contact levels with *f* = 1) produces a difference quickly (10-20 days). Similarly a smaller threshold difference of 5 cases is reached relatively soon. Overall it takes 10-40 days for a substantial difference to arise. The uncertainty in *R*_0_ and other parameters also impacts the time before there is a substantial difference in trajectories (Figure S6). We find that halving uncertainty in the underlying growth rate (by reducing the standard deviation in *R*_0_) can reduce the time until detection from 20-40 days to 12-25 days. Uncertainty in other fixed parameters does not have a strong impact on the time (Figure S6) unless the relaxation conditions are strong (*f*_2_ *>* 0.7). Epidemiological parameters are now somewhat well-established for COVID-19 [26], but we can be less certain of the true value of more behaviour-influenced parameters (*q, u*_*r*_, *u*_d_). We explicitly explore the impact of the incubation period on the time until a substantial difference arises, and find little impact (Figure S7), largely because a few days’ uncertainty in the incubation period is overwhelmed by the the overall time scale and threshold of 10 cases to define a substantial difference. In some instances we cannot differentiate between trajectories for the models with distancing and the models without distancing (these are the missing values in the related figures). Also, if the standard deviation in the underlying parameter *R*_0_ is too large, a substantial difference may not be possible to predict in the time frame of interest.

**Figure 3:**
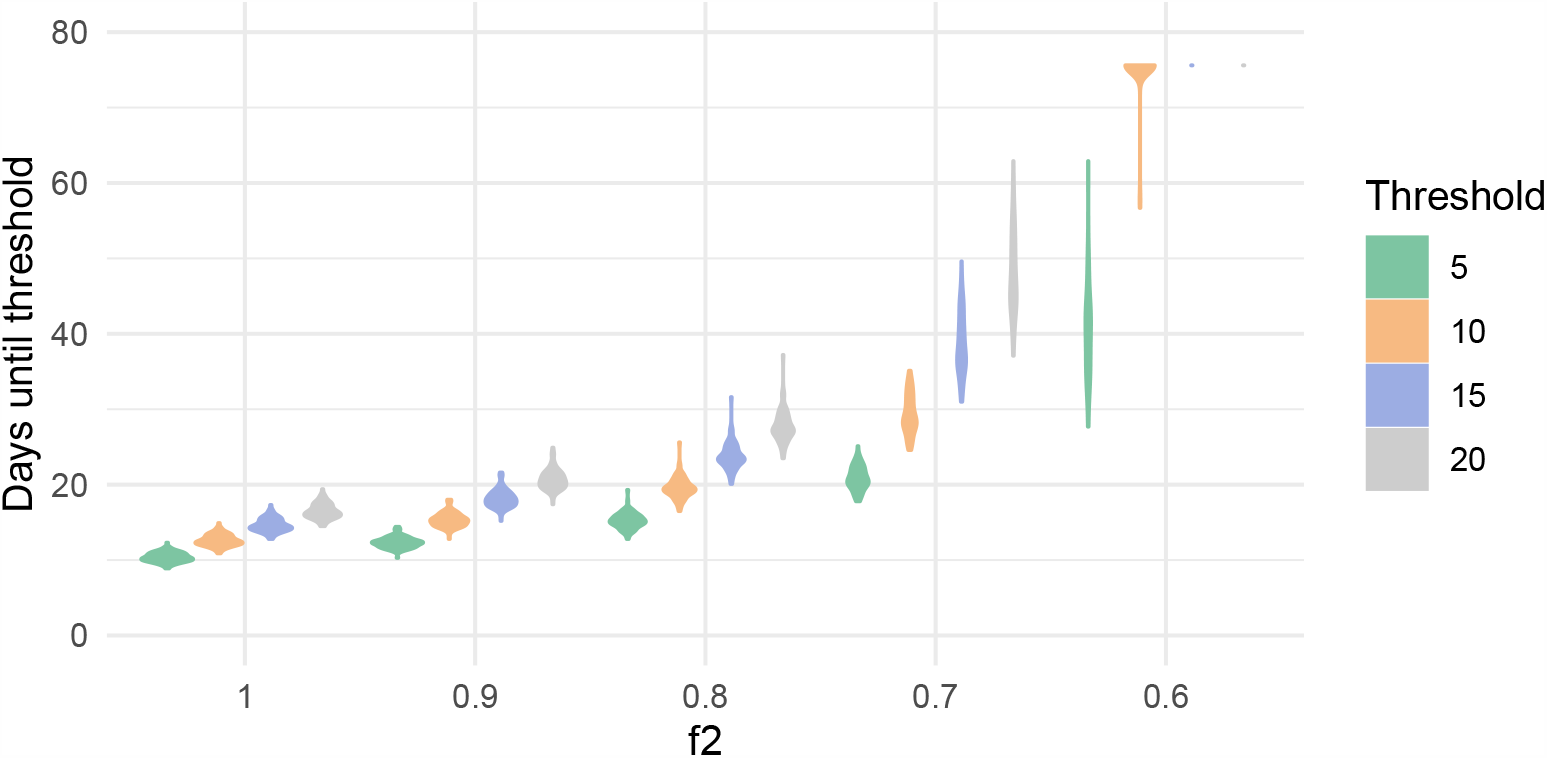
The size of the threshold used to define “substantial differences” impacts the time until such a difference occurs, as does the severity of the change in contact rate *f*. The baseline model has *f*_1_ = 0.36 (on May 17th), and so a change to *f*_2_ = 1 is a complete relaxation of distancing measures, and is detected sooner than a partial relaxation. Each density is produced based on the assumption that *R*_0_ is normally distributed with a mean 2.5 and a standard deviation of 0.15. We vary the threshold for the number of cases of interest in the range {5, 10, 15, 20}. Missing values on the *y*-axis (for example for values of the active cases level of 0.6) indicate that our methods cannot differentiate between the models with and without relaxed physical distancing when the active cases threshold is 15 or 20 cases. For each *f*_2_ and threshold level, we simulate 10,000 ODEs with random initial *R*_0_ values drawn independently from a normal distribution with mean 2.5 and variance 0.15, and for each group of 100 simulated ODEs, we compute the days since May 17th until the excess active cases threshold is detected between the baseline and the condition.

We have conceptually separated the time until there is any substantial difference in model trajectories and the time until we would be able to estimate the impact of a change in distancing, using reported case counts under conditions in British Columbia. These times may differ as a result of reporting delay, but also in some circumstances it may be sufficient to observe signal of a change in distancing under a lower probability or a lower case threshold.

Figure 4 shows the daily maximum likelihood estimate of the physical distancing parameter *f* over time. We find that it took approximately 26 days to accept the value of *f*_1_ (according to the methods described in Section 2.3). We estimate *f*_1_ to be 0.22 on the day of acceptance (April 13th), but this estimate increases to 0.36 by the end of the observation period at 35 days after distancing: April 22nd. Several larger cluster outbreaks had begun to be observed in British Columbia by this time [4], which may have contributed to this increase in the estimated distancing parameter.

**Figure 4:**
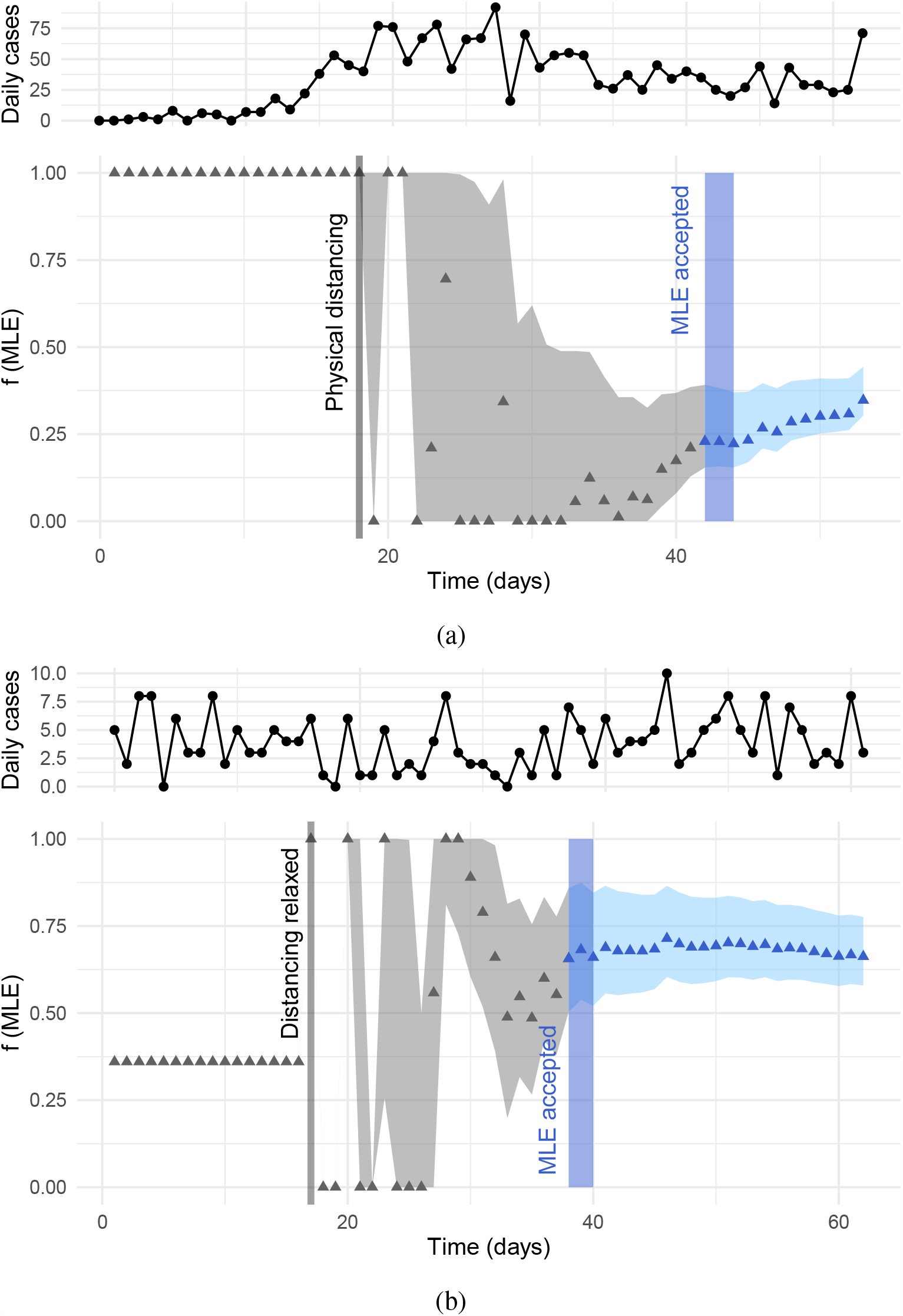
Daily maximum likelihood estimate 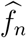 for the strength of social distancing in British Columbia, (a) after implementing distancing, using true observed case counts (26 days to accept MLE) and (b) after relaxing distancing, using simulated data (23 days to accept MLE) under the assumptions that *f* is relaxed to *f*_2_ = 0.65 and the observation noise and delay remain as pre-relaxation during March/April in BC. Grey bands correspond to an estimated 95% credible region, obtained from 50 samples of *R*_0_. The observed (a) and simulated (b) daily case counts are also provided for reference.

In the first few days after the introduction of distancing, we see considerable noise in the daily MLEs. This is expected, as limited data observed in those first few days implies that even small variations in the case counts can have a large effect. The likelihood is flat for a period around the first week after distancing is introduced. The credible intervals are thus more informative than the point estimates. The time at which the credible interval no longer includes 1.0 may indicate the time at which we are confident that distancing has a positive effect (even if we cannot yet determine the size of this effect). We are able to observe this over a much shorter time frame: 8 days until the 95% interval drops below *f*_1_ = 0.99 and 11 days until it drops below *f*_1_ = 0.9, in contrast to the 26 days to accept the MLE estimate. In Figure 4b, we simulate the effects of relaxing physical distancing in British Columbia, from *f*_1_ = 0.36 as estimated in Figure 4a to *f*_2_ = 0.65. Under the assumption that the noise and delay in observing the true outbreak process remains constant, we estimate that it would take 23 days from initiating relaxation of distancing until accepting the MLE 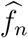. We also performed the same analysis with *f*_2_ = 0.9 and *f*_2_ = 0.5, and estimated 23 days and 30 days respectively. We also used the time-dependent reproductive number [7] (*R*_*t*_) estimates directly derived from reported cases, using an assumed serial interval of 5 (sd 1) days (Figure S8). The time until the 95% quantile for *R*_*t*_ drops below 1.0 is 14 days after distancing was introduced (April 1), and it takes 18, 24 days after distancing is relaxed for *R*_*t*_’s 95% quantile to be above 1.0 (*f*_2_= 0.9, 0.65). Even with weekly smoothing, the *R*_*t*_ values fluctuate greatly, particularly after relaxation of distancing. In contrast, the estimates of *f*_1_ and *f*_2_ do not. However, they are not directly comparable to *R*_*t*_ because we estimate a single *f* value over an extended time period, whereas *R*_*t*_ is a daily (smoothed) value.

The time to observe the strength of distancing is impacted by both the level of noise in the case counts and the delay between symptom onset and reporting. We explore the effects of this in Figure 5. As the noise in the daily case counts is reduced (Figure 5a), so is the time to detect the strength of distancing. Under a level of noise realistic for observed case counts in BC (*ϕ* = 5), we estimated 26 days to accept 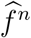. If *ϕ* is doubled (corresponding to halving the variance, approximately, if on the order of 50 cases are observed per day), this reduces to 21 days. Under the most optimistic scenario, where there is no noise (but when there is still an average 8.7 day delay), the time-to-accept is 15 days.

**Figure 5:**
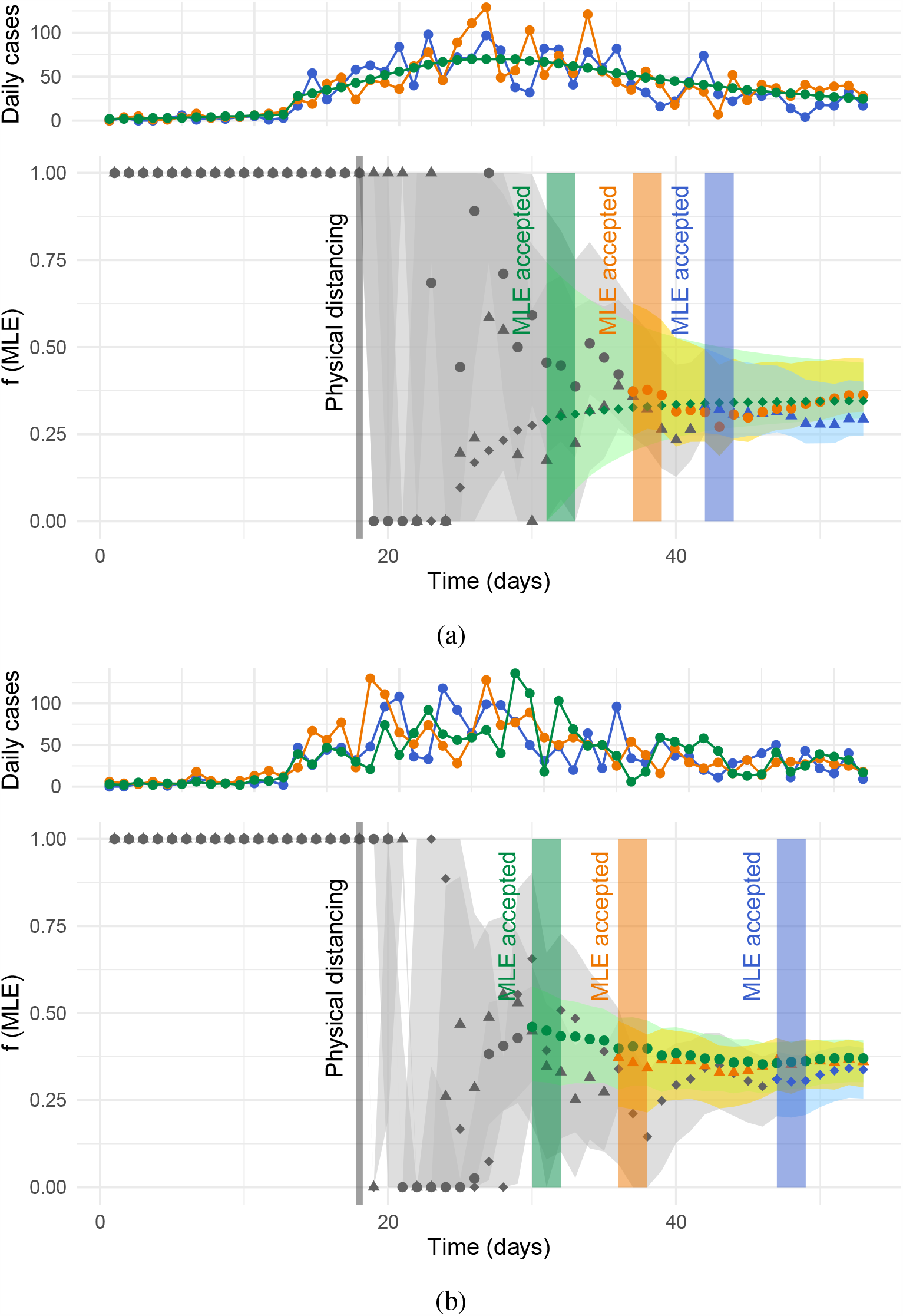
The time to detect the strength of distancing depends on (a) noise in daily case counts and (b) the onset-to-reporting delay. In (a), we vary the dispersion parameter *ϕ* for the daily case counts. Blue: *ϕ* = 5, 26 days to accept MLE. Orange: *ϕ* = 10, 21 days. Green: no noise, 15 days. In (b), we vary the onset-to-reporting delay (mean, variance). Blue: (8.78, 27.4), 32 days to accept. Orange: (4.43, 5.37), 20 days. Green: (0.95, 0.01), 15 days. Physical distancing implemented on day 18 at strength *f*_1_ = 0.36. 95% credible band, produced from 50 samples of *R*_0_ is shown, and MLE accepted window is highlighted with vertical bars. Results are coloured grey before the MLE is accepted. Simulated daily case counts are also provided for reference (top panels).

As would be expected, a longer delay between symptom onset and case reporting also results in a longer time taken to estimate the strength of distancing (Figure 5b). With a Weibull distributed delay of shape 1.73 and scale 9.85 (mean 8.78, variance 27.4) and *ϕ* = 5, as estimated for British Columbia in [2], we estimate 32 days to accept the MLE of *f*_1_. If the delay is reduced to shape 2 and scale 5 (mean 4.43, variance 5.37), the time to accept is 20 days. Lastly, with a delay shape 10 and scale 1 (mean 0.95, variance 0.013), the time to accept is 15 days.

## 4 Discussion

As jurisdictions begin to ease physical distancing measures, we must understand how long it may take to observe a statistically significant difference in reported case counts. We find that it generally takes between 10-60 days before changes in distancing have a substantial, detectable impact on the reported case counts. In certain cases, we find that these methods are unable to differentiate between the relaxed and unrelaxed scenarios; for example, if the deviation in *R*_0_ is high and the relative change in physical distancing is low. Through computing daily estimates of the parameter controlling strength of physical distancing in our model, we find that, at least under public health systems in British Columbia, the time taken to detect changes in distancing is on the order 3-4 weeks. Halving the case count variance or the mean onset-to-reporting delay in British Columbia during March and April could have reduced the time taken to understand the strength of social distancing by approximately 20% and 38%, respectively. This highlights the high potential benefit of improved, consistent surveillance systems, and perhaps contact-tracing apps, which minimise delays and *e*.*g*. weekly patterns or discrepancies in case reporting.

The question of determining when two curves begin to differ has been discussed in general in other literature. Such lines of work include *Simultaneous Equations Models* (SEM), a field within econometrics [25] and growth rates in bacterial culture [16]. These prior works require strong log-linear and normality assumptions and cannot be easily adapted to SEIR-type models. Approaches accounting for the issue of delayed case reporting in epidemiology are often referred to as ‘now-casting’ [21, 33]. These are distinct from the methodology we introduce in this work in that that they focus on inference of the true number of cases at a particular time from the observed number. In contrast, we work backwards from observed cases, using a model which takes observation delay and imperfect sampling into account, to understand the impact of physical distancing on COVID-19.

Our analysis has a number of limitations. Although policy changes happen at defined times, distancing behaviour and other COVID-19 control practices do not change instantaneously, and the time frame to detect changes may therefore be longer than we have estimated. We have not explored a wide range of growth rates or baseline prevalence levels and these may affect the results. The transmission model used is a deterministic SEIR variant, and does not include stochastic effects (except in the observation model), or age or risk structure. The model includes a fraction of the population practising physical distancing, thereby reducing their contact rates, but does not otherwise include heterogeneity in contact patterns. In particular, we focus on the case onset-to-reporting delay, but certain high-risk groups such as healthcare workers may be more likely to get tested and have expedited testing available. Our methodology could readily be extended to structured models, but this requires age-stratified counts and knowledge of the mixing terms. Indeed, any disease model will include exponential growth and decay, and this work is somewhat model-agnostic in that, whatever level of detail goes into producing this exponential behaviour, we can still perform the same eventual inference.

Our approach to determining when the effect of modifying measures is observable relies on using case count data as the indicator for increased community-based transmission. However, public health officials may find outbreaks—even where they do not contribute to statistically higher case counts—by noting epidemiological links among cases (*e*.*g*., links through workplace, family, healthcare or gatherings). Changes in these smaller outbreaks may be detected much faster than our 3 week estimates, but it remains the case that measures directed towards the general population are the main intervention for COVID-19. Our results focus on estimating impacts on this broader population level, and this has long time scales. Sentinel surveillance systems, contact tracing and outbreak detection are among the tools used by public health agencies to gather rapid and more detailed information than case counts during a disease outbreak. These form multi-faceted surveillance networks—including hospitals, primary care, symptom trackers—which may often be faster than confirmed and lab-tested cases. However, these networks also have complex limitations, and greatly vary by jurisdiction. Their data are not always consistently or widely published. Confirmed case counts remain a widely used source for population-level understanding of COVID-19 control, particular for those seeking a broad assessment of COVID-19 in multiple jurisdictions to support policy on borders and travel or to compare effectiveness of control measures.

While we focus on data from British Columbia in this work, our methods may be applied to any region or country with case reporting to determine the relevant time lags and interpret reported cases accordingly. In addition, SEIR-type models are used to forecast COVID-19 in all 50 US states with the *Covid Act Now* project [15]: each state is associated with a COVID-19 risk level based on how soon their projections arrive at certain constant thresholds on measures such as case counts and ICU headroom used. Our methods could improve such region-based work by allowing the parameters and baselines to be calibrated, reflecting population density, testing protocols, demographics and cultural factors regarding social contact. The times to observe the impacts of changes in control measures are likely to be region specific.

We have found in this work that the time-to-detection for a return to widespread transmission owing to relaxed physical distancing measures can be long, indeed considerably longer than than the mean incubation period [26, 14] or often used 14 day time period [5, 27, 17]. In order to decrease the time-to-detection, we need less noisy testing and faster ways to monitor community transmission. Outbreaks within communities contribute both to “noise” in case counts and to uncertainty in *R*_0_, particularly if they reveal areas of previous under-detection. It is therefore also important to maintain consistent surveillance spatially and temporally. Other surveillance techniques could facilitate faster and smoother case detection, but suffer from their own limitations. Consistent sentinel surveillance, *e*.*g*. as are seen in influenza-like-illness data, may have a slightly longer delay, but ultimately less noise. Symptom tracker apps could show changes in incidence sooner than laboratory-confirmed case counts, but could suffer from false positives and may be affected by coverage and usage limitations. Digital contact tracing may also support rapid case finding, but will likely ultimately rely on testing data for confirmation. However, case confirmation delays in a contact tracing context are often considerably shorter than through symptom-based testing [3]. Promising recent research has also investigated detection of SARS-CoV-2 in wastewater [22, 1], which, although potentially not revealing of individual-level infection, may bypass case detection delays and provide an early warning system. Similarly, investigation of live mobility data during the disease outbreak [10, 13] may reveal changes in population behaviour, even if such work requires some assumptions about the link to changes in incidence. Given the long time frames for reported case count data it is urgent that robust combinations of diverse surveillance systems be developed, and for those seeking an overview of COVID-19 trajectories without reference to multi-faceted local surveillance data (perhaps at the national level or to support decisions about travel to and from other jurisdictions), it is important not to over-interpret short-term fluctuations in reported case counts.

## Data Availability

Our data is publicly available from the British Columbia Centre for Disease Control. In addition to that, we've archived a copy of the data on a github site linked in our manuscript.

## Acknowledgements

The work was funded by Genome B.C.’s COVID-19 Rapid Response Funding Initiative (project code COV-142). JES and CC were supported by the federal government of Canada’s Canada 150 research chair program.

## Supplementary Material

**Table S1:**
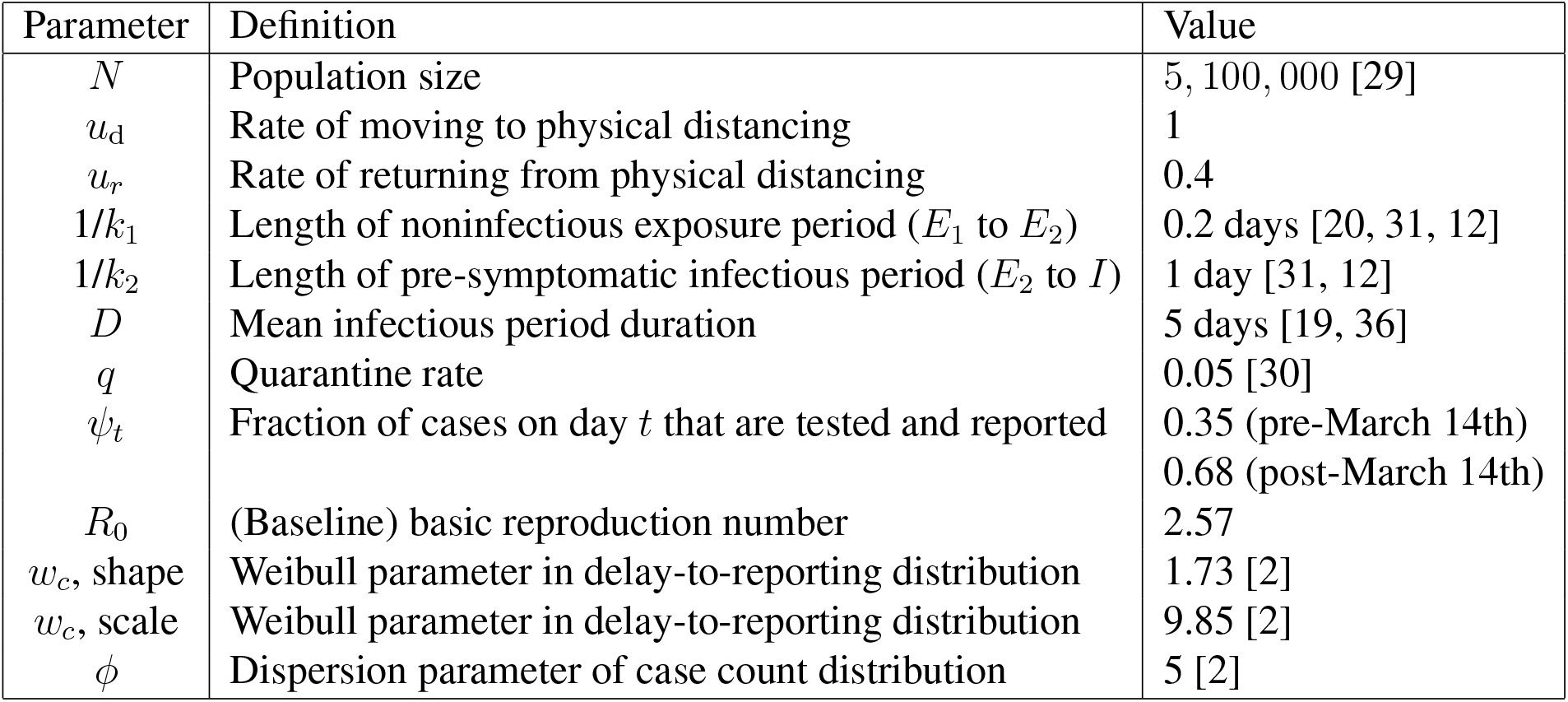
Model parameter values for British Columbia. Values of *ψ*_*t*_ and *R*_0_ are estimated using our model with pre-distancing BC data.

### Impact of uncertainty in model parameters

We consider two experiments wherein the uncertainty about the ODE parameters are modulated, and record the time to detect a threshold of excess in the active cases above baseline models. In our first experiment we focus on *R*_0_. In the main text, we consider ODEs in which the *R*_0_ parameter is initialized to a draw from a normal distribution with mean 2.5 and standard deviation 0.15. This reflects our broad understanding about the uncertainty in *R*_0_. In all conditions, the baseline indicates a situation in which physical distancing is not relaxed, and remains in effect. If the standard deviation is varied away from 0.15, then the days until the threshold is reached (*i*.*e*., a significant difference between the baseline and a relaxed condition is seen) is affected. The results of varying the standard deviation between 0.01 and 0.4 (considering the values in the set {0.01, 0.11, 0.21, 0.31, 0.41}) are shown in Figure S6a. In that Figure, we see that the days until a significant increase in the active cases can vary from 10 to 50 days. Missing values in the lines in this plot indicate that our methods cannot determine whether or not the condition will ever differ from the baseline: if the standard deviation of *R*_0_ is *>* 0.11, we do not predict that a weak relaxation of physical distancing to *f*_2_ = 0.6 will result in a substantial increase in active cases. However, if the standard deviation of *R*_0_ is *<* 0.11 a relaxation of physical distancing to *f*_2_ = 0.6 could lead to a significant increase in active case count after 20 or 40 days.

**Figure S6:**
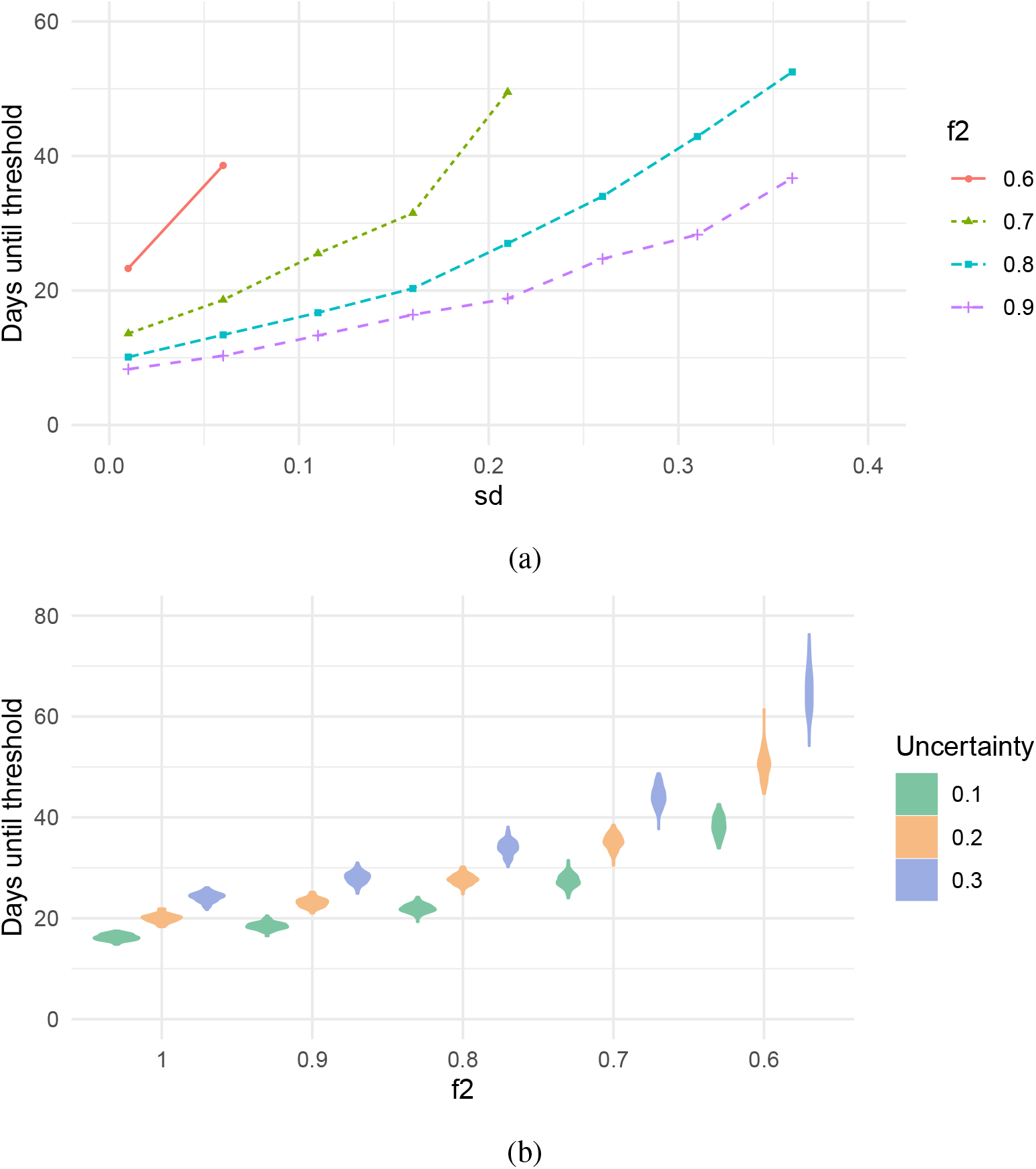
(a) Days until active cases in the relaxed model exceeds the baseline model (y-axis), with varying levels of the standard deviation for *R*_0_ (x-axis). There are four relaxation levels (all starting relaxation on May 17th). Parameter settings are identical to other figures aside from *sd*. The missing values indicate that our methods could not differentiate between the models. (b) The variables *D, k*_1_, *k*_2_, *q, u*_d_ and *u*_*r*_ are all varied between 10% and 30% around their fitted values (with the three levels described in the legend). The value that *f* is relaxed to is shown in the *x*-axis (before relaxation, *f*_1_ is set to 0.36). So, *R*_0_ is sampled at ODE initialization, but so too now are the 6 variables mentioned (with independent draws from the uniform distribution). For each *f*_2_ level, 100 iterations are considered each with 100 replicates.

In another experiment, we consider variation of the ODE parameters *D, k*_1_, *k*_2_, *q, u*_*d*_ and *u*_*r*_. We vary each of these parameters around their default values (given in Table S1) by *X*%, with *X* varied in the set {10, 20, 30}. For each *X* (uncertainty) level, we consider simulations in which the ODE is initialized with random parameters. For the basic reproduction number, *R*_0_ is initialized as a normal distribution with mean 2.5 and variance 0.15 (as is done in the main text), and for the parameters *D, k*_1_, *k*_2_, *q, u*_*d*_ and *u*_*r*_ we initialize each parameter by drawing from a uniform distribution. For each parameter, the range of the uniform distribution has a minimum value of the ‘default’ value of that parameter (given by Table S1) minus *X*% of the default value, and maximum value of the ‘default’ plus *X*%. We then record the days until an active case count excess of 10 is obtained in 95% of the simulations. The number of iterations and the number of replicates is identical to Figure 3 (described in the main text). Density plots for the number of days until the threshold on the excess of active case counts are shown in Figure S6b for a variety of relaxation levels. These results show that if the uncertainty about the parameters of our SEIR-type model are even 10%, then even relaxing distancing totally *f*_2_ = 1 can result in 20 days before the threshold of 10 excess cases is reached. In addition, if social physical is relaxed only moderately (to *f*_2_ = 0.6 for example), then it may be 70 days before an excess of 10 active cases is seen.

**Figure S7:**
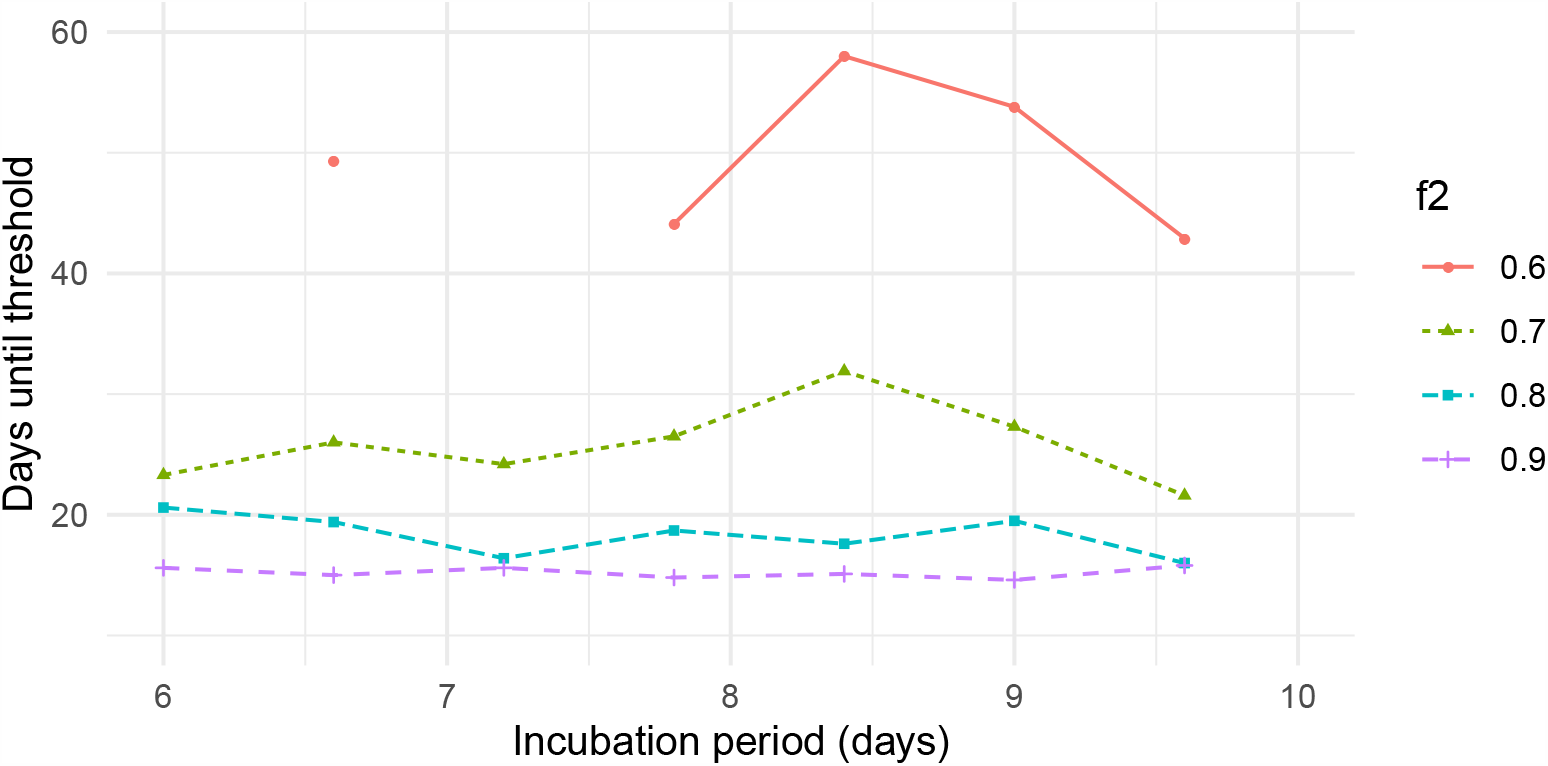
The incubation period has a limited effect on the time before a substantial difference occurs between two simulated scenarios. The initial value *f*_1_ is 0.36. The curves are obtained under the uncertainty assumption that *R*_0_ is normally distributed with a mean 2.5, and a standard deviation of 0.15. The incubation period is 1*/k*_1_ + 1*/k*_2_. For each setting of the incubation period, we rescale *k*_1_ and *k*_2_ evenly from their fitted values (*k*_1_ = 1 and *k*_2_ = 1*/*5) to achieve the incubation period in days. (*e*.*g*., for an incubation period of 7 days 1*/k*_1_ + 1*/k*_2_ = 7, and so the values for *k*_1_ and *k*_2_ used are 1 *∗* (6*/*7) and 1*/*5 (6*/*7) respectively.) Missing values indicate that our methods cannot distinguish between simulations in which physical distancing is relaxed and simulations in which physical distancing is not relaxed (indicating that a relaxation of *f*_2_ to 0.6 may not have an effect for much of the range of incubation periods considered)

The above results indicate that uncertainty about the model parameters substantially increases the number of days until definitive statements about the effects of relaxing social distancing can be made, with the total range of the number of days until effects are seen varying between 20 and 70.

### Effect of the incubation period

We vary the incubation period to explore the impact on the time before the active cases threshold of 10 is reached. The incubation period is defined as 1*/k*_1_ +1*/k*_2_, with default values *k*_1_ = 1*/*5 and *k*_2_ = 1 throughout this work. We vary the incubation period between 6 and 10 days by modulating *k*_1_ and *k*_2_ appropriately (with the same scaling factor for each) and show the results in Figure S7. For each incubation period and relaxation *f*_2_ level, we perform 50 replicates and plot the median number of days until the active cases threshold is reached. We find that there is not much variation in days to threshold as the incubation period is varied. This suggests that the variance in active case due to uncertainty about *R*_0_ may overwhelm variation in the incubation period over the range considered.

### Estimation of *R*_*t*_

For comparison with the daily MLE method applied to British Columbia case count data in Figure 4, we also perform estimation of *R*_*t*_ using *EpiEstim* package [7] in *R*. Results are shown in Figure S8. Note that *EpiEstim* assumes that all cases from the first time step are imported cases, leading to inflated *R*_*t*_ estimates during the first week. To account for this, when finding the the earliest time that *R*_*t*_ is less than/greater than 1.0 at the 95% level, we ignore this first week of estimation.

**Figure S8:**
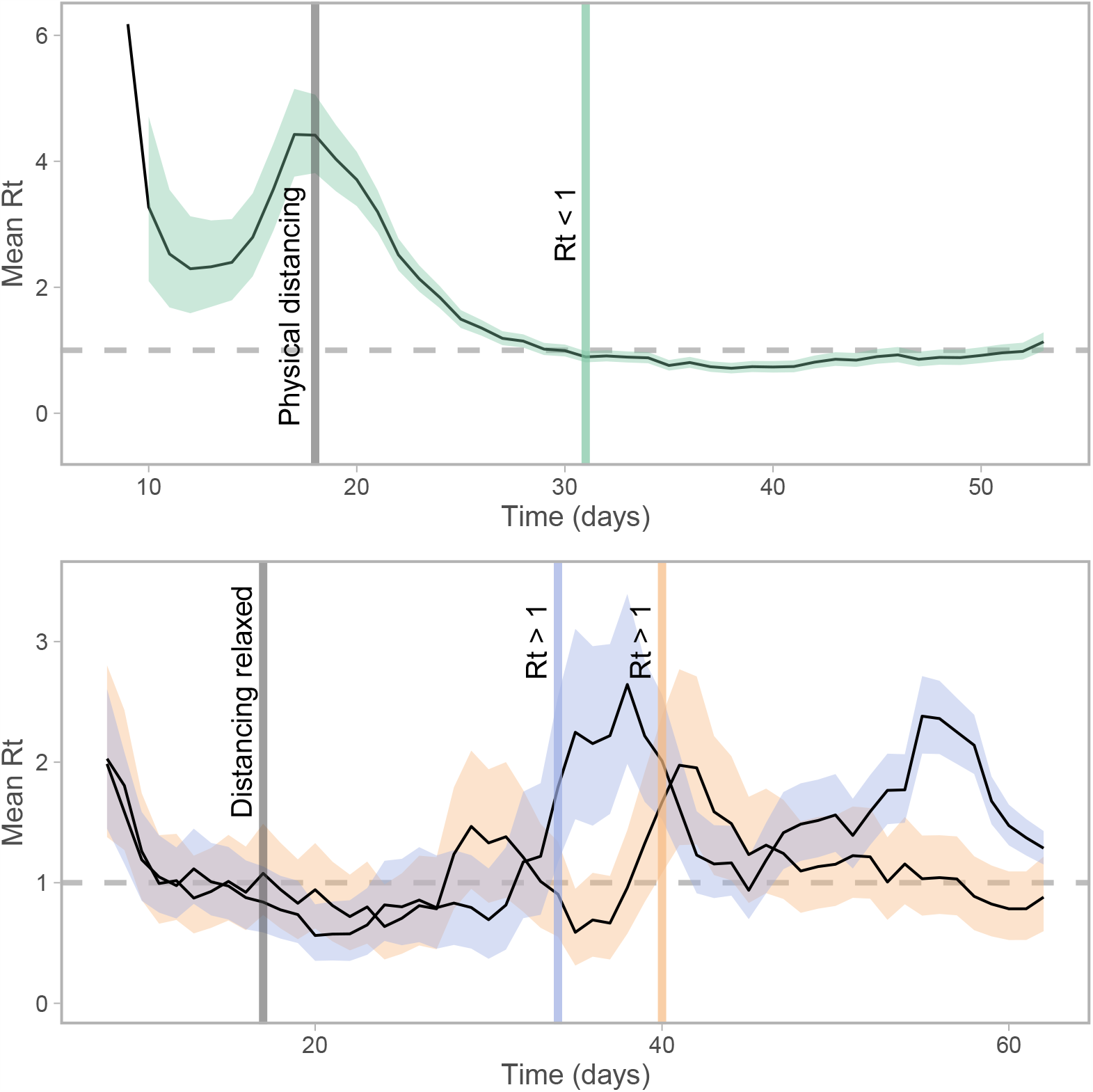
Median estimates of the time-dependent reproduction number *R*_*t*_ in British Columbia, using a weekly sliding window and assumed serial interval of 5 (sd 1) days. Top panel: after implementing distancing, using true observed case counts (14 days until *R*_*t*_ *<* 1 at the 95% level). Bottom panel: after relaxing distancing, using simulated data (18, 25 days until *R*_*t*_ *>* 1 at the 95% level for *f*_2_ = 0.9, 0.65, respectively), under the assumption that *f*_1_ = 0.36 and observation noise and delay remain as pre-relaxation during March/April in BC. Coloured bands correspond to 95% quantiles.

## Footnotes

* http://github.com/carolinecolijn/ObserveEffectsDistancing

